# The burden of vaccine hesitancy for routine immunization in Yaounde-Cameroon: restrictive sampling technique

**DOI:** 10.1101/2022.02.17.22271108

**Authors:** Martin Ndinakie Yakum, Atanga D Funwie, Atem Bethel Ajong, Marcelin Tsafack, Linda Evans Eba Ze, Zahir Shah

## Abstract

Immunization is the most cost-effective health intervention in the world yet, vaccination uptake is still low with less than 50% of children aged 12-23 months fully vaccinated Cameroon. The objective of this study was to estimate the burden of vaccine hesitancy associated with routine EPI vaccines in Yaounde-Cameroon. A two-stage cross-sectional cluster survey was conducted in Yaoundé in May-June 2022, targeting parents/guardians of children 0-59 months. Clusters were selected with probability proportionate to size (PPS) and household’s selection done using a restricted sampling method. Data collection was done using an interviewer-administered questionnaire. Data were cleaned using MS-Excel 2019, and analyzed with R version 4.1.0 (2021-05-18). A total of 529 participants were enrolled out of 708 visited, giving a non-response rate of 25%. In total, vaccine hesitancy was reported in 137(25.90[22.35-29.80] %), and vaccine hesitancy prevalence did not vary significantly across different households’ wealth levels (p-value= 0.3786). However, in wealthy households’ refusal of vaccines (14%) was less than in poorer households (20%). Lack of trust, confidence, and perceived complacency are the leading causes of vaccine hesitancy related to routine immunization in Yaounde-Cameroon. We, therefore, recommend that the burden of vaccine hesitancy should be assessed at national scale and identify sources of misinformation that are at the origin of vaccine hesitancy. Having a clear notion of the effect of social media(Facebook, Instagram, WhatsApp, etc,), radio, TV, and other information sources might guide interventions to combat vaccine hesitancy.

## 1. INTRODUCTION

Vaccination is the most cost-effective public health intervention in terms of the impact on health, society, economies, and education (1). The WHO estimates that [2 to 3 million deaths](http://www.who.int/mediacentre/news/releases/2017/infants-worldwide-vaccinations/en/) are prevented every year through immunization against diphtheria, tetanus, whooping cough, and measles (2).

Despite the merits of vaccination, vaccination coverage is still below the desired level in many places. In 2018, the global immunization coverage varied from 35% for Rotavirus vaccine to 90% for DPT1(first dose of diphtheria, pertussis and tetanus vaccine) (3). Except for a few vaccines like the Rotavirus vaccine, PCV-13 1, and HiB3, the vaccination coverage of all vaccines in the Africa was generally lower than the global coverage (3). According to the results of the 2018 demographic and health survey in Cameroon, approximately 10% of children aged 12-23 months had not received any vaccine dose, and only 41.5% of them had received all the required vaccines (4).

In general, vaccination uptake in a population is determined by vaccination service availability and the level of vaccine hesitancy (5). Concerning vaccination service availability in Cameroon, routine immunization services are available at all public and private health facilities and at all the levels of the health pyramid (6). All services related to routine immunization are free of charge, and integrated health centers (IHCs) organize outreach immunization sessions to cover all remote areas having difficulties reaching the health facility because of distance or/and natural barriers (6). With the level of expansion of the EPI’s activities to all existing health facilities in Cameroon, it is very reasonable to believe that low coverage might be associated more with vaccine hesitancy than its unavailability (7). On the other hand, the situation of vaccine hesitancy is still under-reported(undocumented) in Cameroon.

According to the SAGE Working Group on Vaccine Hesitancy, vaccine hesitancy is defined as the delay in acceptance or refusal of vaccination despite the availability of vaccination services. It is complex and context-specific, varying across time, place and vaccines (5). Vaccine hesitancy is influenced by three (3) key factors related to complacency, confidence, and convenience (5). Several factors contribute to the vaccine decision-making process (8). These factors are the reasons or causes of vaccine hesitancy and are called determinants of vaccine hesitancy. There are two models of vaccine hesitancy determinants which are the 3C model and the 5C model (MacDonald, 2015; Oduwole et al., 2019). The 3C model has to do with 3 determinants (complacency, confidence, and convenience), while the 5C model adds 2 more factors to the three 3C model (rational Calculation and Collective responsibility).

Globally, studies conducted on vaccine hesitancy are still very few, but there is evidence that many countries are struggling with it. More than 90% of the 194 member states of the WHO reported vaccine hesitancy over three years (11,12). The 3-year (2015 to 2017) analysis of the WHO/UNICEF member state Joint Reporting unveiled that vaccine hesitancy is present in all the six WHO regions, and it cuts across all the four categories of country income levels as classified by the WHO (11). In Cameroon, very little is published on vaccine hesitancy; a few that could be identified were related to HPV and covid-19 vaccine and practically nothing on routine EPI vaccines (13–15). Based on these studies, vaccine-related mistrust in Cameroon varied somewhat between 71-85%.

As vaccine hesitancy is multilayered and varies in time, space, and vaccines, it is only normal to have a study on hesitancy related to EPI vaccines to appreciate better the burden of the problem related to routine immunization, and hence contribute to improving vaccine uptake in the country. This study describes the EPI-related burden of vaccine hesitancy in Yaounde, Cameroon.

## 2. MATERIALS AND METHODS

### 2.1. Ethical Approval

This study was approved by the regional ethics committee for the center region of Cameroon (CE No 01410/CRERSHC/2021). Before enrolment, verbal consent was obtained from all participants.

### 2.2. Research design

It was a two-stage cross-sectional community-based cluster survey conducted in Yaoundé in May-June 2022 targeting parents/guardians of children aged 0-59 months. All the six (6) health districts in Yaoundé were included, and clusters were constituted of quarters in the various health area selected with probability proportionate to size (PPS), and household’s selection done using a restricted sampling technique.

Data collection was done using an interviewer-administered questionnaire targeting parents/guardians. This data was cleaned using MS-Excel 2019 and analyzed with R version 4.1.0 (2021-05-18).

### 2.3. Research area

This study was conducted in Yaoundé, the administrative capital of Cameroon. It has a population of more than 2.8 million inhabitants and is made up of six (6) health districts (Biyem-assi, Cite verte, Djoungolo, Efoulan, Nkolbisson, and Nkolndongo).

### 2.4. Study population

This study targeted parents (or guardians) of children aged 0-59 months living in Yaounde during the study period.

### 2.5. Sample size and Sampling technique

The sample size required for this study was estimated at 700 participants. When we considered the feasibility of the work in the field in terms of the number of households that a survey team can complete in one day, we decided to form 30 clusters of 24 households. A total of 30 clusters (quarters) were selected with probability proportionate to size (PPS) using the ENA software version 2021. While in the cluster, the surveyor listed all households with the assistance of a community member (or leader). The direction of sampling in the cluster was selected randomly by tossing a plastic bottle and following the head of the bottle when it stopped rotating. In the same way that we calculate the sampling interval in systematic sampling, the block size(φ) used in the cluster was obtained by dividing the number of households in the cluster by 24 (number of households to interview in each cluster), i.e, N/24 = φ. The surveyor then followed the cluster’s movement plan and randomly selected one (1) household for every successive φ households. In each household selected, parents (preferably the mother) of children aged 0-59 months were interviewed.

### 2.6. Data collection

Data were collected using an electronic questionnaire designed and deployed in tablets with KoBo Toolbox by the investigation team. Prior to data collection, surveyors were recruited and trained on the study procedures, data collection and consenting process for two days. The training of surveyors ended with a pretest of the survey tools, conducted in quarters that were not selected for the survey in Yaounde. Data collection activities in the field were supervised by the principal investigator of the study. Data collections tools used in this study were “Core Closed Questions” and “Likert Scale Questions” proposed by the WHO Vaccine Hesitancy Technical Working Group in 2014(8).

The wealth index is calculated using easy-to-collect data on a household’s ownership of selected assets such as number of people living in one room, type of water source, type of toilet, possession of a television, car, motor bike, telephone, fridge, type of cooking fuel, and type of floor materials for the household wealth index construction.

### 2.7. Data management and data analysis

The database was cleaned using MS-Excel 2019 by visually checking for data consistency. Data analysis was done with R version 4.1.0 (2021-05-18). Principal components analysis (PCA) was used to generate a wealth index.

Vaccine hesitancy was analyzed as proportions of parent’s/guardian’s self-reported vaccine refusal or reluctance in percentage with 95% confidence interval (CI). This was stratified by household wealth level and tested using chi-2 test to appreciate the effect of household wealth on vaccine hesitancy. The threshold of statistical significance was fixed at a p-value <0.05.

Furthermore, the other section of hesitancy was related to scoring some key facts about vaccine and immunization(using Likert Scale Questions), and this was analyzed by calculating the median score for each question and the interquartile range (IQR). It was further compared between different household wealth levels using Mann-Whitney Test to appreciate the effect of household wealth level on vaccine hesitancy. The threshold of statistical significance was fixed at a p-value <0.05.

## 3. RESULTS AND DISCUSSIONS

### 3.1. Sample description

In total, all the 30 clusters planned were reached, and 529 parents were interviewed from 708 households.

Table 1 presents the attitudes and practices of parents vice-a-vice routine immunization services. In total, vaccine hesitancy was reported in 137 participants giving a vaccine hesitancy prevalence of 25.90[22.35-29.80] %. This prevalence did not vary significantly across the different households’ wealth levels (p-value= 0.3786). Approximately 85% of parents believe that vaccination can protect their children from serious illness.

**Table 2:** Self-reported vaccine hesitancy and parents beliefs on childhood vaccines with respect to the various household wealth levels

| Parents believes and aptitudes | Total(N=529) |  | Classes household Wealth Index |  |  |  | OR[95%CI] | p |
| --- | --- | --- | --- | --- | --- | --- | --- | --- |
|  | n | % | I,II&III(N=317) |  | IV&V(N=212) |  |  |  |
|  |  |  | n | % | n | % |  |  |
| Believe that vaccines can protect children from serious diseases | 448 | 84.69 | 26<br>4 | 83.28 | 18<br>4 | 86.79 | 1.32[0.80-2.16] | 0.2717 |
| Think that most parents have their children vaccinated with all the recommended vaccines | 411 | 77.69 | 23<br>7 | 74.76 | 17<br>4 | 82.08 | 1.55[1.00-2.38] | 0.0477* |
| Have been reluctant or hesitated to get a vaccination for child | 115 | 21.74 | 67 | 21.14 | 48 | 22.64 | 1.09[0.72-1.66] | 0.6807 |
| Have ever refused a vaccination for a child | 94 | 17.77 | 64 | 20.19 | 30 | 14.15 | 0.65[0.41-1.05] | 0.0750 |
| Distance, timing of clinic, time needed to get to clinic or wait at clinic and/or costs in getting to clinic prevented me from getting my child immunized | 54 | 10.21 | 36 | 11.36 | 18 | 8.49 | 0.72[0.40-1.31] | 0.2860 |
| Have other pressures in life that prevent from getting child immunized on time | 49 | 9.26 | 35 | 11.04 | 14 | 6.60 | 0.57[0.30-1.09] | 0.0845 |
| There are some reasons to think children should not be vaccinated | 67 | 12.67 | 43 | 13.56 | 24 | 11.32 | 0.81[0.48-1.39] | 0.4470 |
| It is difficult for some ethnic or religious groups in my community / region to get vaccination for their children | 174 | 32.89 | 98 | 30.91 | 76 | 35.85 | 1.25[0.86-1.80] | 0.2365 |
| Have receive or heard negative information about vaccination | 230 | 43.48 | 13<br>1 | 41.32 | 99 | 46.70 | 1.24[0.88-1.77] | 0.2218 |
| Still take my child to get vaccinated after you heard the negative information | 127 | 55.22 | 78 | 59.54 | 49 | 49.49 | 0.67[0.39-1.13] | 0.1292 |
| Religious leaders in my community support vaccines for infants and children | 271 | 51.23 | 15<br>9 | 50.16 | 11<br>2 | 52.83 | 1.11[0.79-1.58] | 0.5468 |
| Political leaders in me community support vaccines for infants and children | 313 | 59.17 | 18<br>2 | 57.41 | 13<br>1 | 61.79 | 1.20[0.84-1.71] | 0.3153 |
| Teachers in my community support vaccines for infants and children | 321 | 60.68 | 18<br>9 | 59.62 | 13<br>2 | 62.26 | 1.12[0.78-1.60] | 0.5420 |
| Health care workers leaders in your community support vaccines for infants and children | 349 | 65.97 | 21<br>1 | 66.56 | 13<br>8 | 65.09 | 0.94[0.65-1.35] | 0.7271 |

In wealthy households, parents reported that most parents vaccinate their children with all vaccine(82%) and less refusal of vaccine(14%) was recorded than in poorer households in which the indicators were reported as 75% and 20%, respectively. Out of 43% of parents that had received negative information on vaccination, only 55% of them still proceeded to vaccinate their children after the information. On the other hand, only 10% reported that distance, timing of clinic, time needed to get to the clinic or wait at the clinic and/or costs in getting to the clinic prevented them from getting their children immunized.

Vaccine hesitancy has been reported to affect all the WHO regions in the world though at different prevalences. In 2016, it was reported that the African region and lower-income countries were the most affected by vaccine hesitancy (16). Another study at household level in Guatemala, Central America, reported a much lower vaccine hesitancy proportion with no vaccine refusal but hesitancy due more to reluctance (17). In Cameroon, a recent study on vaccine hesitancy related to COVID-19 reported a hesitancy proportion of 84.5%(18). Therefore, vaccine hesitancy is present in Cameroon, though it affects EPI vaccines differently from other vaccines.

The very high COVID 19 vaccine hesitancy rate is likely due to its newness and the infodemic associated with the pandemic. Even though EPI vaccine hesitancy in Cameroon seems relatively low, this can be the cause of infant morbi-mortality in Cameroon(2). Table 2 presents EPI vaccines affected by vaccine hesitancy.

**Table 2:** vaccines affected by vaccine hesitancy

| Vaccine | Reluctant/hesitant to receive vaccine for children |  | Refused vaccines for children |  |
| --- | --- | --- | --- | --- |
|  | Freq(n) | Proportion(%) | Freq(n) | Proportion(%) |
| Bcg | 7 | 6.09 | 6 | 6.38 |
| Polio | 41 | 35.65 | 42 | 44.68 |
| Pentavalent Vaccine | 11 | 9.57 | 8 | 8.51 |
| Rotavirus Vaccine | 10 | 8.70 | 7 | 7.45 |
| Measles Vaccine | 15 | 13.04 | 9 | 9.57 |
| Yellow Fever Vaccine | 11 | 9.57 | 6 | 6.38 |
| Pneumococcal Vaccine | 6 | 5.22 | 6 | 6.38 |
| Vitamin A | 9 | 7.83 | 12 | 12.77 |
| Others Vaccines | 42 | 36.52 | 31 | 32.98 |
| All Vaccines | 19 | 16.52 | 14 | 14.89 |

It can be seen that polio is the most affected vaccine having more than 39% hesitancy proportion. For the other vaccines in the EPI, the hesitancy was generally less than 15% as seen on table 2. Vaccine hesitancy varies across different EPI vaccine with the Polio vaccine most affected(see table 3).

**Table 3:** Vaccine hesitancy prevalence by vaccine

| Vaccine | Hesitancy proportion |  |
| --- | --- | --- |
|  | *n* | *Prevalence(%)* |
| Bcg | 9 | 1.70 |
| Polio | 58 | 10.96 |
| Pentavalent Vaccine | 13 | 2.46 |
| Rotavirus Vaccine | 10 | 1.89 |
| PCV-13 | 6 | 1.13 |
| Measles vaccine | 17 | 3.21 |
| Yellow fever vaccine | 12 | 2.27 |
| All vaccines | 22 | 4.16 |

This results are in line with the report of the WHO working group on vaccine hesitancy that describes this phenomenon as “complex”, and varying from one vaccine to the other (8). This could be explained by the polio eradication effort recently in which the Ministry of Health organized countless sessions of polio vaccination campaigns (19,20) over the national territory. This might have raised questions and suspicion within the population. In Northern Nigeria, polio vaccine refusal was reported to be due to lack of confidence, especially because of “too frequent” campaign and due to the the false belief that the Oral Polio Vaccine (OPV) contains birth control ingredients (21). In the same study, the OPV refusal proportion was 33% (21) which is similar to our findings.

According to our findings, causes of vaccine refusal or reluctance were numerous, but the most reported causes included the respondent had heard or read negative information about vaccine on social media (>40%), the respondent did not think that the vaccine was needed (>29%), bad experience with previous vaccination (>13%), and bad experience with a vaccinator in the past (>8%). In any case, our findings show that lack of trust and confidence, perceived Complacency and Convenience are the leading causes of vaccine hesitancy in Yaounde-Cameroon (see figures 1 & 2).

**Figure 1:**
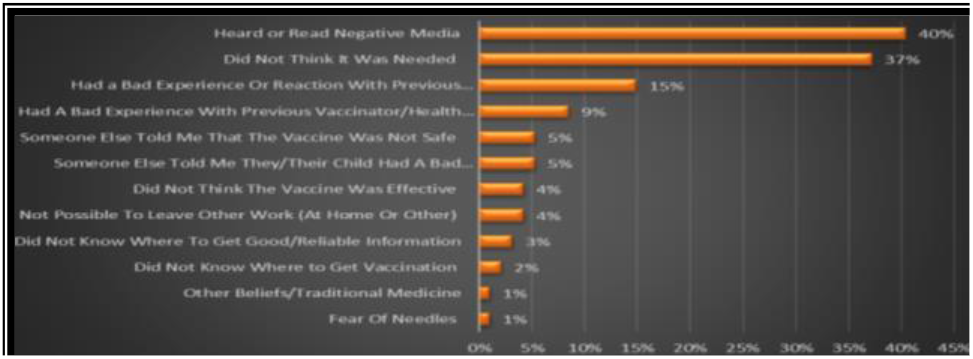
Reasons stated by the parents for refusing one or more vaccines for their children

**Figure 2:**
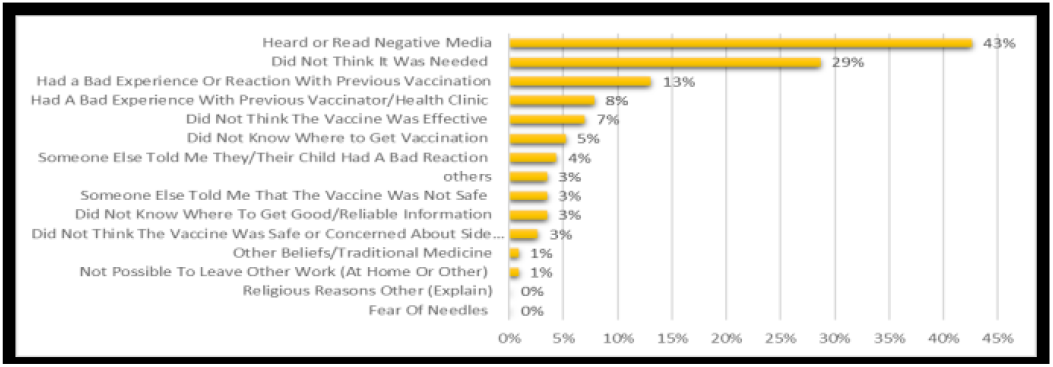
Reasons stated by the parents for being reluctant to accept one or more vaccines for their children

This suggest that lack of trust and confidence is the leading cause of vaccine hesitancy related to routine immunization in Yaounde-Cameroon. Nothwithstanding, Convenience equally plays a significant role as shown in figures 1&2. These results are in accordance with the report of the WHO Working group on vaccine hesitancy and other publications that stated that Lack of confidence, Complacency and Convenienceare the three main factors that cause vaccine hesitancy (8,9,22–25).

Vaccine hesitancy has a very significant impact on the health of individuals and the general public health of their surroundings. When vaccine hesitancy occurs, children immunization is either delayed or refused (8) leading to reduced effectiveness of immunization interventions. The unvaccinated child is exposed to infectious diseases and possible death (26). At the same time, the unimmunized child is a major threat to other children around them as they can easily transmit inoculums capable of causing outbreaks and deaths (26,27) and consuming the already limited health resource. In a similar way, vaccines already in stock are not used on time, leading to a waste an additional waste of the limited human, financial and material resources.(28).

Targeting vaccine hesitancy for routine immunization in Yaound-Cameroon should be a major priority of the EPI. If awareness is raised on the importance of immunization and finding a way to combat misinformation on media, it will go a long way to improve routine immunization uptake in Yaounde-Cameroon.

Table 4 presents the Likert survey questionnaire presented by vaccine hesitancy. It suggests that the level of knowledge and the perceptions of vaccine-hesitant and non-hesitant parents/guardians on immunization are significantly different.

**Table 4:** Parents Opinion on vaccination in children

| Statements | Vaccine hesitancy |  |  |
| --- | --- | --- | --- |
|  | Present Median (IQ) | Absent Median (IQ) | P |
| Childhood vaccines important for my child's health | 5.0(2.0) | 5.0(0.0) | 0.0000* |
| Childhood vaccines are effective | 5.0(2.0) | 5.0(0.0) | 0.0000* |
| Having my child vaccinated is important for the health of others in my community | 5.0(2.0) | 5.0(1.0) | 0.0000* |
| All childhood vaccines offered by the government program in my community are beneficial. | 5.0(2.0) | 5.0(1.0) | 0.0005* |
| New vaccines carry more risks than older vaccines | 5.0(1.0) | 5.0(2.0) | 0.0323* |
| The information I receive about vaccines from the vaccine program is reliable and trustworthy. | 4.0(3.0) | 5.0(2.0) | 0.0002* |
| Getting vaccines is a good way to protect my child/children from disease. | 4.0(2.0) | 5.0(2.0) | 0.0026* |
| Generally, I do what my doctor or health care provider recommends about vaccines for my child/children. | 4.0(2.0) | 4.0(2.0) | 0.0006* |
| I am concerned about serious adverse effects of vaccines. | 5.0(1.0) | 5.0(2.0) | 0.1003 |
| My child/children do or do not need vaccines for diseases that are not common anymore. | 3.0(4.0) | 3.0(2.5) | 0.0204* |

Our findings are in line with findings from previous studies in which vaccine hesitancy was found to be caused by beliefs and lack of knowledge on immunization (17,21). Another study in India reported vaccine hesitancy clustering on social media(29). This further supports the fact that improving awareness and finding a way to target social media misinformation might be a way to combat vaccine hesitancy in Cameroon.

## CONCLUSIONS

Routine EPI Vaccine hesitancy in Yaounde is 25.90%, and there was no statistical significance in vaccine hesitancy proportion accross different households’ wealth levels. In wealthy households’ refusal of vaccines (14%) was less than in poorer households (20%). The lack of trust and confidence and perceived complacency are the leading causes of vaccine hesitancy related to routine immunization in Yaounde-Cameroon.

There is a need for the public health authorities in Yaounde and Cameroon as a whole to design interventions to minimize routine immunization hesitancy thereby improve immunization coverage for EPI. In particular, fighting rumors and force information and educating mothers on the importance of immunization might go a long way to reduce vaccine hesitancy in Yaounde. We, therefore, recommend that the burden of vaccine hesitancy be assessed at national scale and the sources of misinformation causing vaccine hesitancy clearly identified and controlled. Having a clear notion of the effect of social media (Facebook, Instagram, WhatsApp, etc,), radio, TV, and other information sources can better guide interventions to combat their contribution to vaccine hesitancy.

## Data Availability

All data produced in the present study are available upon reasonable request to the authors

## ACKNOWLEDGEMENTS

We will like to thank data collectors namely: Miss DOUANLA KOUTIO Ingrid Marcelle, Miss TCHENGO MASSOM THÉRÈSE ZITA, Miss Christelle Bertyl TCHANA MBETBEUM, and Miss Ngueni Letegnou Nancy. We are very thankful to them for their total commitment and professionalism permitted timely completion of data collection in the field while ensuring optimal data quality for the project. We are equally very thankful to Professor Djuidje Marceline for supporting us in the preparation for the data collection phase of the project. She provided us with a space and other training materials in her laboratory at the University of Yaounde I, where the data collection teams were trained before being deployed to the field.

